# Side by side comparison of three fully automated SARS-CoV-2 antibody assays with a focus on specificity

**DOI:** 10.1101/2020.06.04.20117911

**Authors:** Thomas Perkmann, Nicole Perkmann-Nagele, Marie-Kathrin Breyer, Robab Breyer-Kohansal, Otto C Burghuber, Sylvia Hartl, Daniel Aletaha, Daniela Sieghart, Peter Quehenberger, Rodrig Marculescu, Patrick Mucher, Robert Strassl, Oswald F Wagner, Christoph J Binder, Helmuth Haslacher

## Abstract

**Background:** In the context of the COVID-19 pandemic, numerous new serological test systems for the detection of anti-SARS-CoV-2 antibodies have become available quickly. However, the clinical performance of many of them is still insufficiently described. Therefore we compared three commercial, CE-marked, SARS-CoV-2 antibody assays side by side.

**Methods:** We included a total of 1,154 specimens from pre-COVID-19 times and 65 samples from COVID-19 patients (≥14 days after symptom onset) to evaluate the test performance of SARS-CoV-2 serological assays by Abbott, Roche, and DiaSorin.

**Results:** All three assays presented with high specificities: 99.2% (98.6-99.7) for Abbott, 99.7% (99.2-100.0) for Roche, and 98.3% (97.3-98.9) for DiaSorin. In contrast to the manufacturers’ specifications, sensitivities only ranged from 83.1% to 89.2%. Although the three methods were in good agreement (Cohen’s Kappa 0.71-0.87), McNemar’s test revealed significant differences between results obtained from Roche and DiaSorin. However, at low seroprevalences, the minor differences in specificity resulted in profound discrepancies of positive predictability at 1% seroprevalence: 52.3% (36.2-67.9), 77.6% (52.8-91.5), and 32.6% (23.6-43.1) for Abbott, Roche, and DiaSorin, respectively.

**Conclusion:** We find diagnostically relevant differences in specificities for the anti-SARS-CoV-2 antibody assays by Abbott, Roche, and DiaSorin that have a significant impact on the positive predictability of these tests.

## Introduction

COVID-19 is a new disease caused by Severe Acute Respiratory Syndrome Coronavirus 2 (SARS-CoV-2), which was first described by Chinese scientists in early January 2020 (1). On March 11, the WHO officially declared the novel SARS-CoV-2 infections a pandemic, which has now spread rapidly across the entire globe, with almost 6.5 million confirmed cases and over 375,000 confirmed deaths (2). COVID-19 is characterized by a broad spectrum of individual disease courses, ranging from asymptomatic infections to the most severe cases requiring intensive medical care (3). The reliable detection of infected persons and, subsequently, their isolation is essential for the effort to prevent the spread of the SARS-CoV-2 virus quickly and efficiently. Therefore, reverse transcriptase-polymerase chain reaction (RT-PCR) testing is required for direct detection of the pathogen. Unfortunately, RT-PCR testing does not always give a clear answer to whether the SARS-CoV-2 infection is currently present or not (4,5). On the other hand, serological testing for SARS-CoV-2 specific antibodies can be used as an additional diagnostic tool in case of suspected false-negative RT-PCR results (6) or for individual determination of antibody levels. Moreover, cross-sectional serological studies provide essential epidemiological information to allow a correct estimation of the spread of the disease within a population (7,8). The first commercially available serological SARS-CoV-2 tests, mostly standard ELISA tests or lateral flow rapid tests, have not always proved to be sufficiently specific and sensitive (9,10). Recently, the first tests for fully automated large-scale laboratory analyzers have been launched. The present evaluation aims to compare three of these test systems manufactured by Abbott (11), Roche (12), and DiaSorin (13), with particular emphasis on specificity, which is crucial for an adequate positive predictive value given the current low seroprevalence worldwide.

## Materials and methods

### Study design and patient cohorts

This nonblinded prospective study aims at a detailed comparison of three automated SARS-CoV-2 detection methods with a particular focus on specificity and positive predictability. A total of 1,154 samples from three cohorts of patients/participants with sampling dates before 01.01.2020 were used to test specificity. The samples derived from three different collections: a cross-section of the Viennese population, LEAD study (14), preselected for samples collected between November and April to enrich seasonal infections (n=494); a collection of healthy voluntary donors (n=302; 269 individuals, 11 donors with a 4-fold repetition of the donation within a median period of 4.5 years [3.6-5.5]); a disease-specific collection of samples from patients with rheumatic diseases (n=358).

For estimation of test sensitivity, samples of 65 COVID-19 donors/patients with a symptom onset to analysis time of ≥14 days (median time interval of 41 [28-49] days) were evaluated in parallel on all three analysis platforms. For asymptomatic donors (n=6), SARS-CoV-2 RT-PCR confirmation to analysis time was used instead. We subjected only a single serum sample per patient to sensitivity analysis to avoid data bias due to uncontrolled multiple measurement points of individual patients.

Supplementary Table 1 gives a comprehensive overview of characteristics and cohort-specific inclusion and exclusion criteria; Supplementary Table 2, 3, and Supplementary Fig. 1 provide additional descriptive statistics on donors/patients included in the cohorts. All included participants gave written informed consent for donating their samples for scientific purposes. From patients, only left-over material from diagnostic procedures was used. The overall evaluation plan conformed with the Declaration of Helsinki as well as with relevant regulatory requirements. It was reviewed and approved by the ethics committee of the Medical University of Vienna (1424/2020).

### Biomaterials

Used serum samples were either left-over materials from diagnostic procedures (Department of Laboratory Medicine, Medical University of Vienna) or part of a sample cohort processed and stored by the MedUni Wien Biobank. All pre-analytical processes were carried out according to standard operating procedures in an ISO 9001:2008/2015-certified (MedUni Wien Biobank, Department of Laboratory Medicine) and ISO 15189:2012-accredited (Department of Laboratory Medicine) environment. Standard sample protocols were described previously (15).

### Antibody testing

SARS-CoV-2 specific antibodies were measured according to the manufacturers’ instructions on three different automated platforms at the Department of Laboratory Medicine of the Medical University of Vienna.

1. IgG antibodies against SARS-CoV-2 nucleocapsid (SARS-CoV-2 IgG) were quantified employing a chemiluminescent microparticle immunoassay (CMIA) on the Abbott ARCHITECT® i2000sr platform (Abbott Laboratories, Chicago, USA). For the cut-off of ≥1.4 Index (S/C), the manufacturer gives a negative percentage agreement (NPA, corresponding to specificity) of 99.60% (95% CI: 98.98 – 99.89) calculated from samples collected before the COVID-19 outbreak and of 100.00% (95.07 – 100.00) in patients with other respiratory illnesses. Regarding diagnostic sensitivity, 0.00% (0.00 – 60.24) are reported <3 days after symptom onset, 25.00% (3.19 – 65.09) on days 3 – 7, 86.36% (65.09 – 97.09) on days 8 – 13, and 100.00% (95.89 – 100.00) from day 14 on (11). According to the manufacturer, the assay is designed for the qualitative detection of IgG antibodies to SARS-CoV-2.
2. The Elecsys® Anti-SARS-CoV-2 assay (Roche Diagnostics, Rotkreuz, Switzerland) was applied on a Cobas e 801 modular analyzer. It detects total antibodies against the SARS-CoV-2 nucleocapsid (N) antigen in a sandwich electrochemiluminescence assay (ECLIA). For the suggested cut-off of ≥1 COI, the manufacturer reports specificities of 99.80% (95% CI: 99.58 – 99.92) for samples derived from diagnostic routine, 99.83% (99.51 – 99.97) for blood donors, and 100% (91.19 – 100) for both a common cold panel and a Coronavirus panel. Sensitivity was estimated as 65.5% (56.1 – 74.1) during days 0 – 6 post RT-PCR confirmation, 88.1% (77.1 – 95.1) from day 7 to day 13, and 100% (88.1 – 100%) from day 14 on (12). According to the manufacturer, the system delivers qualitative results, either being reactive or non-reactive for anti-SARS-CoV-2 antibodies.
3. The LIAISON® SARS-CoV-2 S1/S2 IgG test detects IgG-antibodies against the S1/S2 domains of the virus’ spike protein in a chemiluminescence immunoassay (CLIA). The test was applied to a LIAISON® XL Analyzer (DiaSorin S.p.A., Saluggia, Italy). The manufacturer reports a diagnostic specificity of 98.5% (95% CI: 97.5 – 99.2) in blood donors and 98.9% in presumably SARS-CoV-2 negative diagnostic routine samples (94.0 – 99.8). Applying a cut-off >15.0 AU/mL (borderline results 12.0 – 15.0, require a re-test algorithm), the test’s sensitivity is reported time-dependently with 25.0% (14.6 – 39.4) ≤5 days after RT-PCR-confirmed diagnosis, 90.4% (79.4 – 95.8) from day 5 to day 15, and 97.4% (86.8 – 99.5) after >15 days (13). Samples that repeatedly tested borderline were classified as positive. The manufacturer indicates to provide quantitative measurement results on the system.

### Statistical analysis

Unless stated otherwise, continuous data are given as median (quartile 1 – quartile 3). Categorical data are given as counts and percentages. Diagnostic sensitivity and specificity, as well as positive and negative predictive values, were calculated using MedCalc software 19.2.1 (MedCalc Ltd., Ostend, Belgium). 95% confidence intervals (CI) for sensitivity and specificity were calculated according to Clopper and Pearson (“exact” method) with standard logit confidence intervals for the predictive values (16). Receiver-Operating-Characteristic (ROC)-curve analysis was used to evaluate test accuracy and compare the diagnostic performance of the three test systems, according to DeLong et al. (17). Between-test agreements were assessed by interpretation of Cohen’s Kappa-statistics, and further evaluated with McNemar’s tests. Statistical significance was assumed at p<0.05. Figures were produced with MedCalc software 19.2.1 and GraphPad Prism 8 (GraphPad Software, San Diego, USA).

## Results

### Specificity

To describe assay specificity, we used a total of 1,154 serum samples collected before SARS-CoV-2 circulated in the population and which are, by definition, negative for SARS-CoV-2 specific antibodies. The three different specificity cohorts A-C (described in detail in Supplementary Table 1-3 and Supplementary Figure 1) presented with different rates of false-positives (Table 1) - cohort C (cohort of rheumatic diseases) showing the highest reactivities. We found in total 9, 3, and 20 false-positive samples for Abbott, Roche, and DiaSorin, leading to an assay specificity of 99.2 (95%CI: 98.6-99.7), 99.7% (95%CI: 99.2-100.0), and 98.3% (95%CI: 97.3-98.9) respectively (Figure 1A-C). Median and 90th percentile values of negative samples were 0.025 and 0.115 Index for Abbott, 0.0815 and 0.0927 COI for Roche, and below LOD and 5.52 AU/ml for DiaSorin. False-positive samples yielded median values of 2.21 Index (2.14-2.67) for Abbott SARS-CoV-2 IgG (cut-off: ≥1.4 Index), 1.65 COI (1.47-1.72) for Roche Elecsys® Anti-SARS-CoV-2 (cut-off: ≥1 COI), and 22.4 AU/ml (17.38-57.35) for DiaSorin LIAISON® SARS-CoV-2 S1/S2 IgG (negative <12.0 AU/ml, equivocal 12.0 AU/ml <= x <15.0 AU/ml, positive ≥15.0 AU/ml).

**Table 1.**
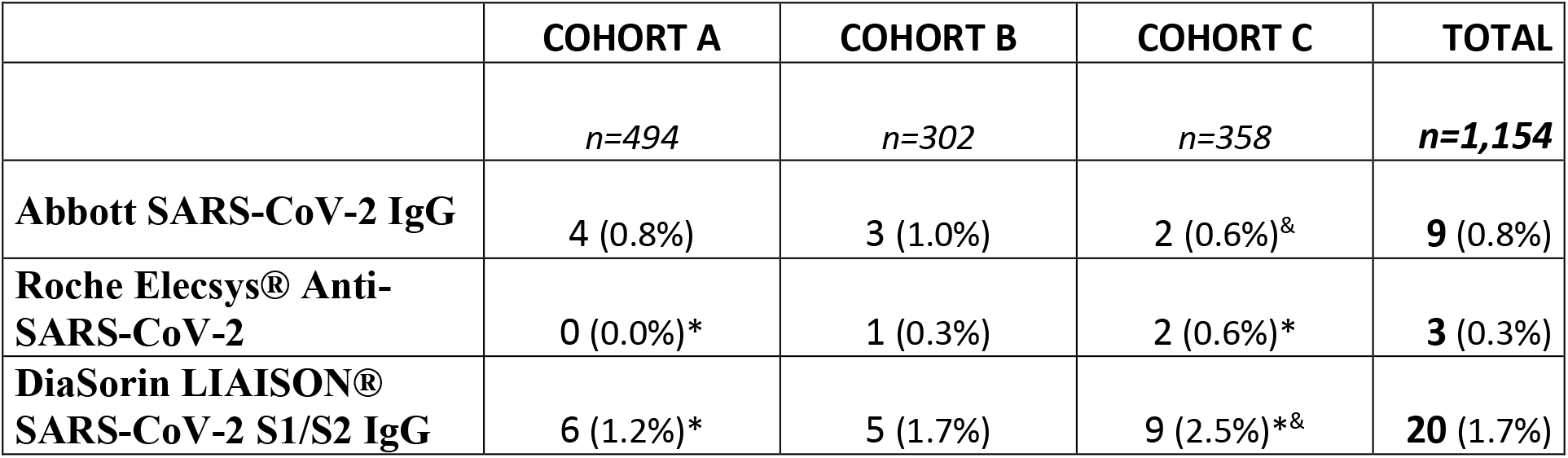
Numbers and percentages of false positive SARS-CoV-2 antibody reactivities in three different specificity cohorts: Cohort A (LEAD-Study), Cohort B (Healthy donor collective), and Cohort C (Rheumatic diseases cohort). X^2^-tests for differences of proportions: ^*^… Roche vs. DiaSorin p<0.05, ^&^… Abbott vs. Diasorin p<0.05

|  | COHORT A | COHORT B | COHORT C | TOTAL |
| --- | --- | --- | --- | --- |
|  | *n=494* | *n=302* | *n=358* | ***n=1,154*** |
| **Abbott SARS-CoV-2 IgG** | 4 (0.8%) | 3 (1.0%) | 2 (0.6%)<sup>&</sup> | **9 (0.8%)** |
| **Roche Elecsys® Anti-SARS-CoV-2** | 0 (0.0%)* | 1 (0.3%) | 2 (0.6%)* | **3 (0.3%)** |
| **DiaSorin LIAISON® SARS-CoV-2 S1/S2 IgG** | 6 (1.2%)* | 5 (1.7%) | 9 (2.5%)*<sup>&</sup> | **20 (1.7%)** |

**Figure 1:**
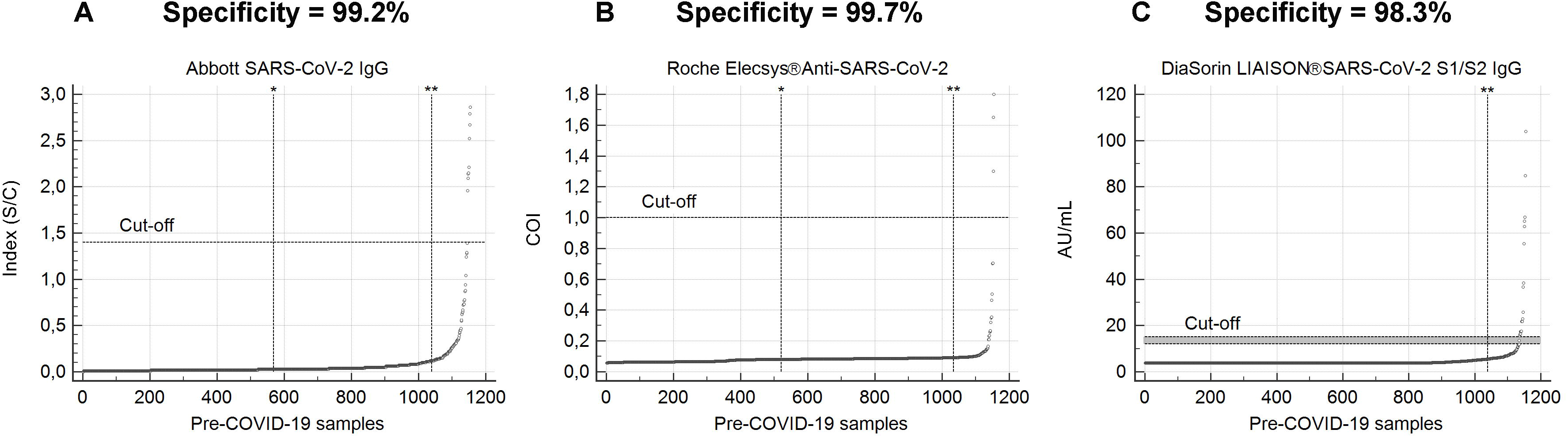
Specificity was determined using 1,154 serum samples taken before the circulation of SARS-CoV-2. For SARS-CoV-2 antibody tests Abbott SARS-CoV-2 IgG (**A**), Roche Elecsys® Anti-SARS-CoV-2 (**B**), and DiaSorin LIAISON® SARS-CoV-2 S1/S2 IgG (**C**), values of specificity samples are shown in rank order. Horizontal dotted lines mark the respective cut-offs recommended by the manufacturer and, in the case of DiaSorin, a gray zone for equivocal results. Vertical dotted lines indicate the median (^*^) and the 90th percentile values (^**^). Median and 90th percentile values of negative samples were 0.025 and 0.115 for Abbott, 0.0815 and 0.0927 for Roche, below LOD and 5.52 for DiaSorin.

### Sensitivity

To estimate assay sensitivity, we used serum samples from 65 donors/patients at later time points following SARS-CoV-2 infection, at least ≥14 days after symptom onset (median interval of 41 [28-49] days). In this late phase, we assumed the majority of donors/patients having reached prominent and constant levels of SARS-CoV-2 specific antibodies. Surprisingly, we could find a relatively high percentage of samples that were testing negative for SARS-CoV-2 antibodies: Abbott 10, Roche 7, and DiaSorin 11 false-negatives leading to calculated sensitivities of 84.6% (73.6-92.4), 89.2% (79.1-95.6), and 83.1 (71.3-91.2)%, respectively. Five serum samples were consistently tested negative in all three assays despite being derived from individuals tested positive for SARS-CoV-2 by RT-PCR. All seven false-negatives in the Roche test overlapped with false-negatives in the Abbott test (both nucleocapsid-antigen based assays), whereas DiaSorin was negative for an additional six serum samples exclusively (S1/S2-domain antigen-based assay).

### PPV and NPV

Although specificity and sensitivity are essential criteria for assessing the quality of a test procedure, they have little informative value about the probability of a positive/negative test result, indicating the presence/absence of SARS-CoV-2 specific antibodies without taking prevalence into account. Therefore, a comparative overview for specificity, sensitivity, as well as positive and negative predictive values at 1%, 5%, and 10% SARS-CoV-2 antibody seroprevalence is shown in Figures 2A and 2B and summarized in Table 2. While the differences between the test systems for varying seroprevalences do not have a significant impact on NPV (range 98.1%-99.9%), the consequences for PPV are pronounced. At seroprevalence rates of 10%, all three systems show acceptable PPVs of 92.3.%, 97.4%, and 84.2% for Abbott, Roche, and DiaSorin, but at 1% seroprevalence these drop to unsatisfactorily or even unacceptably low values of 52.3% (36.2-67.9), 77.6% (52.8-91.5), and 32.6% (23.6-43.1) for Abbott, Roche, and DiaSorin.

**Figure 2:**
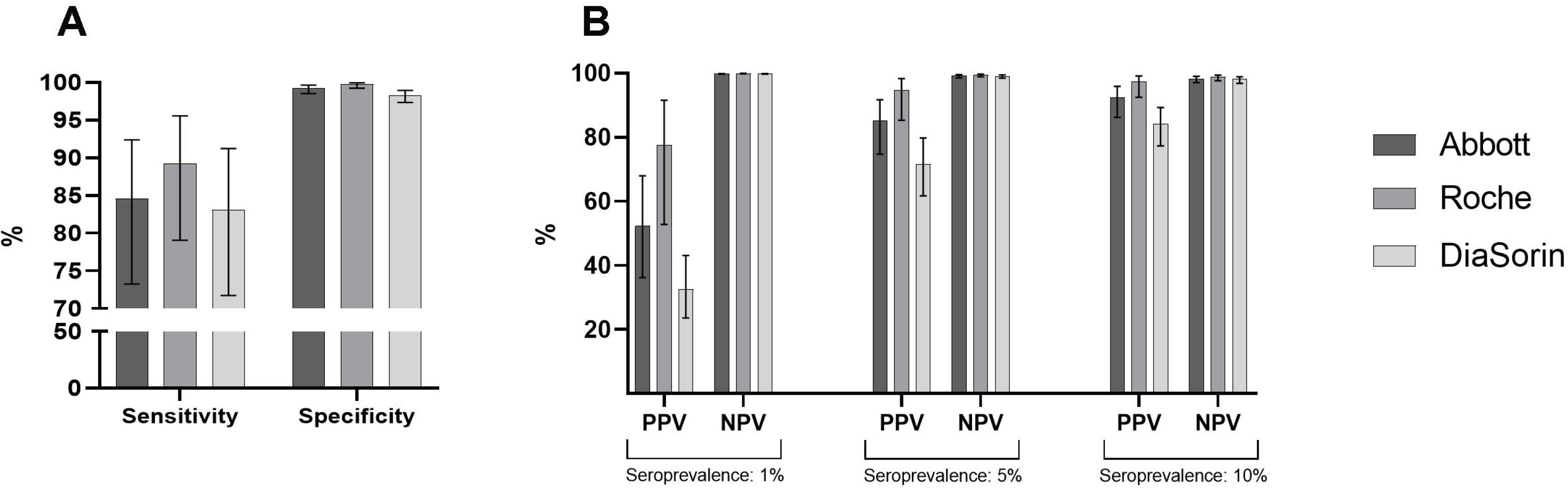
Quality criteria of the SARS-CoV-2 antibody tests Abbott SARS-CoV-2 IgG, Roche Elecsys® Anti-SARS-CoV-2, and DiaSorin LIAISON® SARS-CoV-2 S1/S2 IgG. Columns represent values of sensitivity, specificity (**A**), PPV, and NPV (**B**) at 1%, 5%, and 10% assumed seroprevalence; bars indicate the 95% CI.

**Table 2.**
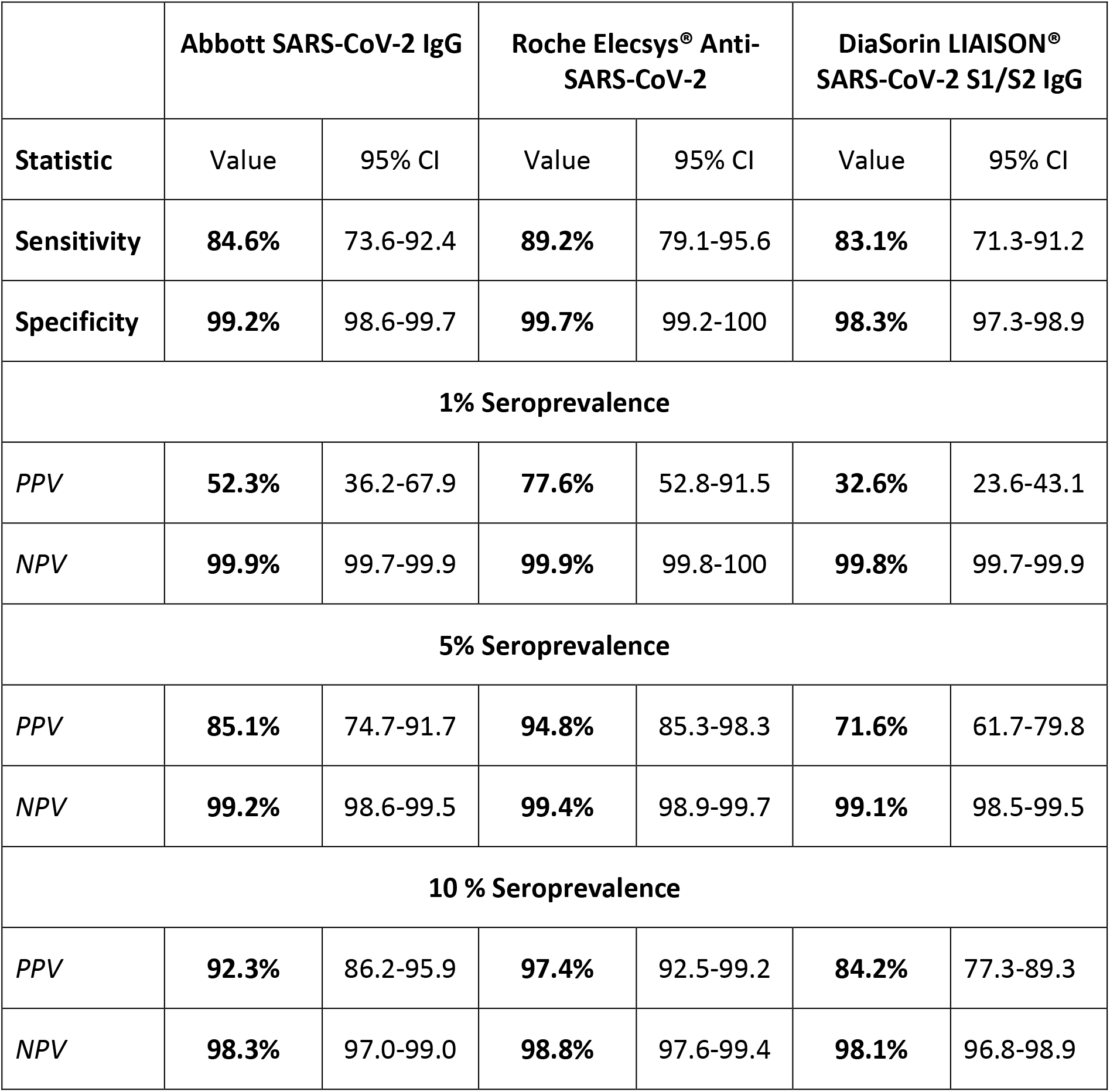
Values for Specificity, Sensitivity, Positive-Predictive-Value (PPV) and Negative-Predictive-Value (NPV) at 1%, 5% and 10% SARS-CoV-2 seroprevalence (SP) with 95% confidence intervals (95% CI).

### ROC Curve Analysis

As shown in Figure 3A-C, all three ROC curves presented with areas under the curves (AUC) above 0.97 (Abbott: 0.994 [95% CI: 0.987-0.997], Roche: 0.989 [0.981-0.994], DiaSorin: 0.977 [0.967-0.985]). Comparison of ROC-AUCs, according to DeLong et al., did not reveal significant differences (Differences: Abbott/Roche p=0.487, Abbott/DiaSorin p=0.112, Roche/DiaSorin p=0.395). In the next step, we aimed to assess whether modifying the cut-off values could improve the explanatory power of the ROC-curves.

**Figure 3:**
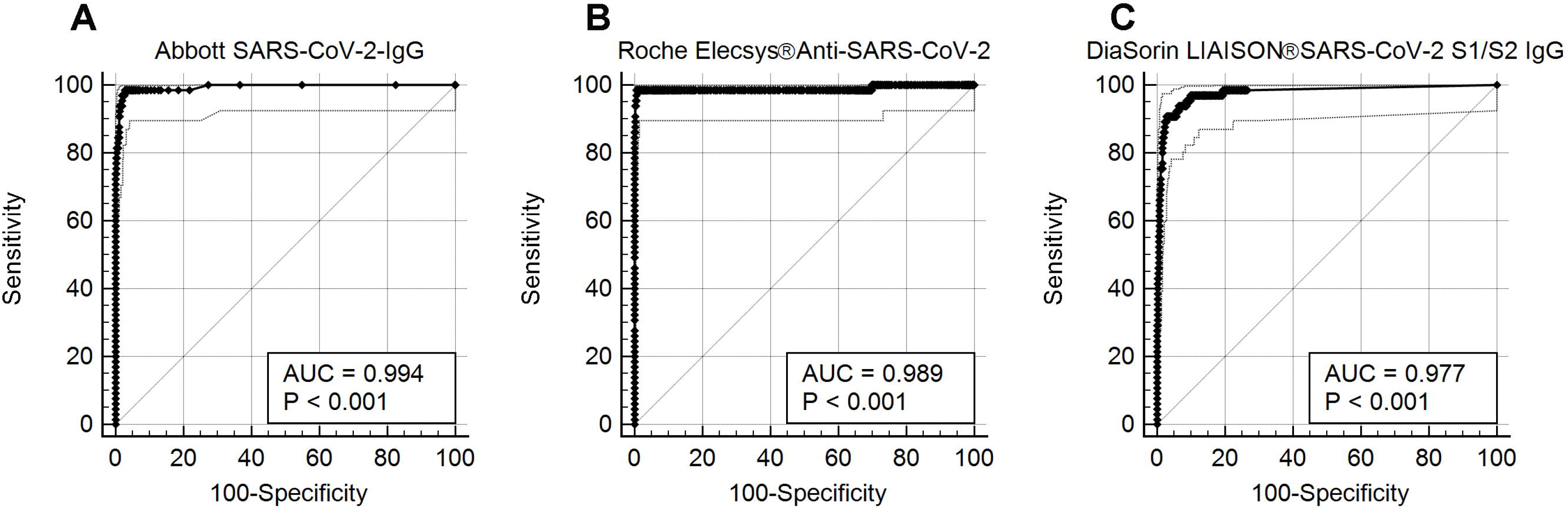
Receiver Operating Characteristic (ROC) curves for Abbott (**A**), Roche (**B**), and DiaSorin (**C**) are shown. The Area Under the Curve (AUC) indicates the test accuracy, 95% confidence intervals are represented by gray dotted lines.

Cut-offs associated with the Youden’s index (maximum sum of sensitivity and specificity) of >0.42, >0.355, and >8.76 for Abbott, Roche, and DiaSorin lowered the PPV considerably, being as low as 26.4 (20.3-33.6), 62.1 (43.9-77.4), and 24.8 (18.9-31.9) at 1% seroprevalence for Abbott, Roche, and DiaSorin (Supplementary Table 4).

### Between-test agreement/disagreement

Correlation analysis of measurement values between the different platforms showed only moderate to weak concordance. The Pearson correlation coefficient was r=0.66 (p<0.001), for both Abbott/DiaSorin (both IgG-assays) and Abbott/Roche (both nucleocapsid-antigen based assays). In contrast, Roche/DiaSorin, with a coefficient of r=0.25 and a hardly reached significance of 0.044, could only show a very weak correlation.

Therefore, the test systems’ agreements were studied in a pairwise fashion applying inter-rater agreement statistics (Cohen’s Kappa). The agreement between Abbott and Roche was very good (0.87 [0.81-0.94]). Agreement between Abbott and DiaSorin, and DiaSorin and Roche was good: 0.71 (0.62-0.80), and 0.76 (0.67-0.84), respectively (Table 3). Despite a good overall inter-rater agreement, significant differences could be shown using McNemar’s test for DiaSorin and Roche (Supplementary Table 5).

**Table 3.** Inter-rater agreement (Cohen’s kappa) with linear weights. Value of K <0.20 poor agreement, 0.21-0.40 fair agreement, 0.41-0.60 moderate agreement, 0.61-0.80 good agreement, and 0.81-1.00 very good agreement.

|  | Abbott |  |  | Kappa (95% CI) |
| --- | --- | --- | --- | --- |
| Roche | NEG | POS |  | **0.87 (0.81-0.94)** |
| NEG | 1149 | 9 | 1158 (95.0%) | Standard Error |
| POS | 6 | 55 | 61 (5.0%) | 0.032 |
|  | 1155 (94.7%) | 64 (5.3%) | 1219 |  |
| DiaSorin | NEG | POS |  | **0.71 (0.62-0.79)** |
| NEG | 1131 | 14 | 1145 (93.9%) | Standard Error |
| POS | 24 | 50 | 74 (6.1%) | 0.045 |
|  | 1155 (94.7%) | 64 (5.3%) | 1219 |  |

|  | DiaSorin |  |  | Kappa (95% CI) |
| --- | --- | --- | --- | --- |
| Roche | NEG | POS |  | **0.76 (0.67-0.84)** |
| NEG | 1136 | 22 | 1158 (95.0%) | Standard Error |
| POS | 9 | 52 | 61 (5.0%) | 0.042 |
|  | 1145 (93.9%) | 74 (6.1%) |  |  |

## Discussion

To the best of our knowledge, this is the first side-by-side comparison of three fully automated SARS-CoV-2 antibody tests applying more than 1,200 distinct donor/patient samples. We identified significant differences between two of the three systems, especially regarding positive predictability at the expectable low prevalence rates. SARS-CoV-2 is a new virus closely related to the betacoronaviruses SARS-CoV and MERS. Like SARS-CoV-2, those highly virulent pathogens cause severe respiratory syndromes, often with lethal outcome (18). In contrast, infections with other members of the coronavirus family usually present with mild colds, including 229E, OC43, NL63, and HKU1 (19). Compared to SARS-CoV (which is no longer circulating), cross-reactivity between SARS-CoV-2 and endemic seasonal coronaviruses is low. To date, with few exceptions (24), no accumulation of cross-reactivities between anti-SARS-CoV-2 antibodies and seasonal coronavirus antibodies has been found. We have therefore refrained from screening a coronavirus panel for possible cross-reactivity.

### Specificity

To best describe the specificity of a serological test, it is essential to have a reliable reference, i.e., to ensure that the samples used are negative for the target analyte. For SARS-CoV-2, this means using serum/plasma samples obtained before the first appearance of the new virus. Therefore, we have compiled large pre-COVID-19 cohorts, which have the following characteristics: A) samples of an age and sex-controlled population-based cohort of more than 11,000 participants (LEAD-Study) (14), randomly chosen from Vienna and surrounding areas (n=494). B) samples of healthy voluntary donors (n=302), which are typically used at our Department for the evaluation of new assays, and C) samples of a disease-specific collection of patients with rheumatic diseases including rheumatoid arthritis and systemic lupus erythematodes (n=358), known to have a high prevalence of autoantibodies and other atypical immune activities, enhancing the potential of interference with serological testing. We found several false-positives in the rheumatological cohort (n=13), and to a lesser extent in the other two cohorts (n=9 in the healthy donor cohort and n=10 in the LEAD study). Notably, an overlap of samples tested false-positive in the different systems did not typically occur, and only one of 32 samples tested positive in more than one assay (Abbott and DiaSorin, one sample from the LEAD study). Since these two test systems use different antigens (nucleocapsid vs. S1/S2 proteins) but the same detection method (IgG), this false-positive reaction is likely associated with interference of the IgG measurement. Calculated specificities are strongly dependent on the spectrum and the size of a selected specificity cohort. If we calculated the specificities of each cohort separately, we would be able to report variable specificities: cohort A (Abbott 99.2%, Roche 100%, DiaSorin 98.8%), cohort B (Abbott 99%, Roche 99.7%, DiaSorin 98.3%), and cohort C (Abbott 99.4%, Roche 99.4%, DiaSorin 97.5%). Roche would range from ideal 100% down to 99.4%, the same level as the best result for Abbott, and DiaSorin would be nearly as good as the worst Abbott specificity or show a 2.5% difference to the best Roche value. This would have an enormous impact on prevalence dependent parameters like PPV. A recent evaluation of the DiaSorin LIAISON® SARS-CoV-2 S1/S2 IgG assay with 1,140 pre-COVID-19 samples reported a specificity of 98.5% (20), nearly perfectly matching the specificity of 98.3% we found when calculating the average of all three cohorts. In contrast, another recent study reported a specificity of 100% for DiaSorin. However, the authors used only n=81 samples for specificity testing (21). Similarly, a further evaluation comparing all three SARS-CoV-2 tests by Abbott, Roche, and DiaSorin found quite different specificities, namely 100%, 98%, and 96.9% for Abbott, Roche, and DiaSorin, respectively. Again, the specificity cohort was very small (n=100, and n=98 for DiaSorin) (22). This underlines the importance of selecting adequately sized testing cohorts to obtain reliable and comparable results. In summary, the specificities of 99.2%, 99.7%, and 98.3% found in the present study are very close to the values given by the manufacturers of 99.6%, 99.8%, and 98.5% for Abbott, Roche, and Diasorin, which were also established on large collectives.

### Sensitivity

The COVID-19 positive cohort used in this study for the estimation of sensitivities is relatively small (n=65). However, it has three distinctive features:

1. each patient/donor is represented in the collective with only one serum sample, avoiding bias of the data by multiple measurements of the same individuals,
2. the median time of blood sampling was 41 days after onset of symptoms and thus in the plateau phase of antibody formation, and
3. 80% of the cohort were non-hospitalized COVID-19 patients (two-thirds of them with mild symptoms), and only 20% were intensive care patients.

As sensitivity within the first 14 days after symptom onset is highly variable for most SARS-CoV-2 antibody assays but becomes better >14 days (23,24), we expected high sensitivities for all tested assays in the plateau phase of antibody formation. Surprisingly we found multiple RT-PCR confirmed COVID-19 patients displaying very low antibody titers that did not surpass the respective assay-specific cut-offs and therefore were considered negative. Five samples were negative in all three assays: all were RT-PCR confirmed cases, 4/5 non hospitalized (42-51 days after symptom onset), two with mild and the other two with moderate symptoms, and symptom duration of <1 week for all. None of these patients had a known immune dysfunction or other severe diseases. One patient was an ICU patient with an underlying hematological disease, and the sample was taken at day 15 after symptom onset. This patient mounted a partial antibody response starting from day 21 after symptom onset becoming positive in the Abbott assay (2.21 Index) and reaching positivity on day 30 also in the DiaSorin assay (29.1 AU/ml). At this late point, Abbott measured 4.19 Index, whereas Roche remained (since day 21) at a level around 0.2 COI. This example illustrates that although the vast majority of patients show compatible and plausible results in different test systems, single cases can display a complex picture of time-courses and reactivities in specific assay systems that are still poorly understood. Another interesting observation was that in six patients with positive detection of SARS-CoV-2 specific antibodies in the Roche and Abbott test, DiaSorin failed to detect antibodies. This observation, combined with the claim that the detection of S1/S2 protein-specific antibodies is equivalent to the detection of neutralizing antibodies (nAbs) (20), raises the fundamental question of whether nAbs are detectable in all patients with confirmed COVID-19 infection. There is evidence that neither the assumption of equivalence of nAbs and S1/S2 protein-specific antibodies (25) nor the assumption that all COVID-19 patients produce measurable titers of nAbs is universally valid (26). A clear answer to the question of whether antibody measurements against nucleocapsid- or spike protein-associated antigens are more sensitive and specific, and how these behave in relation to nAbs assays (the postulated gold standard in terms of sensitivity and specificity) is not possible based on the data currently available.

### PPV, NPV, ROC-Analysis, test agreement

Specificity and sensitivity alone are not sufficient to judge the performance of a diagnostic test; prevalence-dependent accuracy measures like PPV and NPV are necessary, and especially PPV, in times of low prevalence (27). For most regions affected by the pandemic, the prevalence of SARS-CoV-2 antibody-positive individuals is unknown but can be estimated to be below 5%. Therefore, for all SARS-CoV-2 EUA approved antibody tests, the FDA compares the performance of the assays based on a 5% seroprevalence (28). At this rate, the results presented here show PPV values of 85.1% (74.7-91.7), 94.8% (85-98), and 71.6% (61.7-79.8) for Abbott, Roche, and DiaSorin, respectively. The PPV values between Roche and DiaSorin differ so clearly that not even the 95% CI intervals overlap. Therefore, we must assume that these two assays differ significantly from each other in terms of positive predictability. Using these two tests at lower seroprevalences, such as 1%, leads to an even more pronounced difference between Roche and DiaSorin (77.6% vs. 32.6%) and an unacceptable low PPV of 32.6% (23.6-43.1) for DiaSorin.

Although the area under the curves (0.994, 0.989, and 0.977 for Abbott, Roche, and DiaSorin) did not differ significantly from each other (sensitivity cohort size was too small), modeling of the cut-offs according to Youden’s index revealed interesting insights: only Roche could increase the sensitivity without losing specificity dramatically (Sensitivity: 89.2% ➔ 98.5%; Specificity: 99.7% ➔ 99.4%; cut-off: >0.355 COI). In contrast, DiaSorin at the suggested cut-off of >8.76 AU/ml (similar to (20)) increased the sensitivity from 83.1% to 90.8% but worsened the specificity from 98.3 to 97.2%. In line with this, despite a good overall agreement between Roche and DiaSorin results (Cohen’s Kappa 0.76 [0.67-0.84]), the McNemar’s test still showed significant differences, indicating disagreement (in particular in false-positives) more often than expected by chance.

The strength of this study is the side by side evaluation of three assays with a large number of negative samples to give reliable and comparable specificity data (no missing data). Limitations are the moderate numbers of positive samples. Moreover, obtained sensitivities cannot easily be compared to other studies because of the unique feature of our COVID-19 cohort, including 80% non-hospitalized patients with mainly mild symptoms. The latter is highly relevant for a potential use of antibody tests to assess seroprevalence in large populations.

## Conclusion

We find diagnostically relevant differences in specificities for the anti-SARS-CoV-2 antibody assays by Abbott, Roche, and DiaSorin that have a significant impact on the positive predictability of these tests. We conclude that low seroprevalences require an unusually high specificity for SARS-CoV-2 antibody tests, which pushes some test systems to their limits earlier than others. Therefore, the choice of the test must depend on the respective seroprevalence, and strategies such as confirmation of possible false-positive test results with additional testing must be considered.

## Data Availability

Data will be made freely available after peer-review. Until this, data can be requested from the corresponding author.

## Acknowledgements

We sincerely thank Marika Gerdov, Susanne Keim, Karin Mildner, Elisabeth Ponweiser, Manuela Repl, Ilse Steiner, Christine Thun, Martina Trella, for excellent technical assistance. Finally, we want to thank all the donors of the various study cohorts. Without their voluntary participation in the establishment of the biobanks, this study would not have been possible.

**List of abbrevations**
COVID-19: Coronavirus disease 2019
SARS-CoV-2: Severe Acute Respiratory Syndrome Coronavirus 2
RT-PCR: Reverse transcriptase-polymerase chain reaction
ECLIA: Electrochemiluminescence assay
95% CI: 95% confidence interval
CMIA: Chemiluminescent microparticle immunoassay
NPA: Negative percentage agreement
CLIA: Chemiluminescence immunoassay
ROC: Receiver-operating-characteristics
AUC, AUROC: Area under the (ROC-)curve
LOD: Limit of detection
PPV: Positive predictive value
NPV: Negative predictive value
nAbs: Neutralizing antibodies
EUA: Emergency Use Authorization

